# Operationalizing multimorbidity in relation to mortality and physical function: findings from a prospective longitudinal cohort study

**DOI:** 10.1101/2024.07.22.24310803

**Authors:** Han Sang Seo, Daniel Davis, Amal R. Khanolkar, Alun Hughes, Praveetha Patalay

## Abstract

**BACKGROUND AND OBJECTIVES:** There is little consensus on how multimorbidity should be operationalized in life course research. We set out to derive better empirical definitions for multimorbidity by examining its different operationalizations and their associations with mortality, functional independence and physical capability in a longitudinal population-representative cohort.

**RESEARCH DESIGN AND METHODS:** We used data from 2653 (51.6% female) study members in the MRC National Survey of Health and Development (NSHD), a British cohort born in 1946. We examined predictive utility of five multimorbidity operationalizations by age 63 (*binary multimorbidity, unweighted disease count, weighted disease count, clustered multimorbidity, cumulative multimorbidity*) using 16 chronic disease, to predict 12-year mortality and age 69 functional independence and physical capability (grip strength, chair rise speed, balance).

**RESULTS:** The multimorbidity operationalizations of *unweighted disease count* (16.3%) and *weighted disease count* (16.7%) explained the highest variation for mortality compared with other operationalizations. Similarly, explained variation for *unweighted disease count* (17.0%) and *weighted disease count* (16.4%) showed the highest prediction for functional independence and physical capability at age 69 years, with minor variations in physical performance measures where clustered multimorbidity also explained high variance (e.g. 8.6% for changes in chair rise speed). Binary multimorbidity, although frequently used in research was the least predictive of all outcomes.

**DISCUSSION AND IMPLICATIONS:** The associations between various multimorbidity measures and mortality and physical functioning lend support to important influence of multimorbidity on later-life health and functioning while highlighting the differences in variance prediction and point estimates when different approaches to operationalizing multimorbidity are used. An unweighted disease count approach might be suitable for many epidemiological research questions as it is simple to estimate while being as predictive as other more complex approaches such as weighted disease counts.

**Translational Significance:** Multimorbidity research urgently needs a standardized approach to its measurement and operationalization. This study simultaneously compares different strengths of association between multimorbidity definitions and mortality, functional independence and physical capability. This adds possibility by using the predictive utility for mortality, functional independence and physical capability as criteria for determining the usefulness of a given operationalization. Quantifying how these vary offers practical choices between methods of operationalizing multimorbidity for various purposes in research and clinical settings, for example, a targeted strategy aimed at understanding mortality, physical function and health care utilization.

## Background and Objectives

Multimorbidity is a public health priority for health and social care practitioners, researchers, and policy-makers.^1^ More than half of older people aged 65 or over are living with multimorbidity, the prevalence of which is projected to increase from 54% to 68% over next two decades.^2^ With the progress of medical science and increasing life expectancy, the prevalence of multimorbidity is increasing worldwide.^3^ It is expected to further increase in the coming years, now becoming the norm in the elderly.^4^ The prevalence of multimorbidity tends to be higher in older adults as people become more susceptible to chronic illnesses with increasing age.^5^

Multimorbidity has been associated with multiple undesirable outcomes including higher mortality,^6^ disability^7^ and lower functional capacity,^8^ decreased quality of life,^9^ polypharmacy and increased treatment burden.^12^ Moreover, earlier age at multimorbidity onset prolongs the period spent in poorer health thereby widening socioeconomic and health inequalities over the life course.^12^

Several studies have attempted to identify specific multimorbidity patterns in older adults, beyond determining its basic presence or absence. Studies consistently show that chronic diseases tend to cluster in specific patterns. Moreover, multimorbidity may have a differential health impact if its constituent conditions are disparate or closely related.^16^ Individuals with disparate conditions may require more complex care, as long-term conditions in different physiological systems are more likely to result in polypharmacy and result in competing medical demands, in contrast to conditions within the same system.^13^

The wide variation in multimorbidity prevalence, from 13.1% to 71.8% in population studies^15^ arise because there is no consensus on its conceptual or operational definitions. This includes counting the number and types of chronic conditions, thresholds, data sources, study populations and cohort characteristics such as age, sex and socioeconomic position. Estimates are higher if more chronic conditions are included, or if definitions are broader.^18^

Individuals with multimorbidity generally experience a higher risk of mortality in comparison to those without.^7^ The biological plausibility linking multimorbidity and mortality mirrors physiological mechanisms that heighten the risk of death in people with diseases. In a meta-analysis of 26 studies by Nunes et al.^6^ involving participants aged over 65 years, the presence of ≥2 or ≥3 diseases was linked to a pooled relative hazard ratio for mortality of 1.73 and 2.72, respectively. The mortality rate, on average, saw a 40% increase with the addition of each extra disease. Furthermore, mortality in older people is multifactorial and includes environmental and socioeconomic characteristics,^21^ as well as being influenced by geriatric conditions ^22^ and healthcare actions. ^23^

Multimorbidity has been associated with greater functional limitations and a decline in physical functioning. Limitations in physical function due to multimorbidity decisively affect people’s illness and treatment burden and their response capacity, which may further increase multimorbidity.^24^ Apart from a few exceptions,^25^ the association between multimorbidity and poor physical function in older adults was demonstrated in several cross-sectional and longitudinal studies.^26–32^ According to a population-based study by Aarts et al., functional impairment from multimorbidity persists over time and often shows further worsening.^27^ Jindai et al. showed that, in older individuals, the association between multimorbidity and functional limitation is strengthened in older age (>75) and females.^28^ Measures of physical function, being important biomarkers of older adults’ health statuses, can be used as an alternative to disease status for assessing the health of older persons.^31^

We aimed to derive better empirical definitions for multimorbidity by examining associations with later-life physical function and mortality. This endeavour aimed to offer insights useful for enhancing healthcare delivery and formulating health policies tailored to individuals affected by multimorbidity. It adds to the literature by conducting a population-based analysis that involves study members being followed up throughout their lives. In particular, we measured the predictive utility of different multimorbidity constructs including *binary multimorbidity, unweighted disease count, weighted disease count, clustered multimorbidity,* and *cumulative multimorbidity* based on the duration of conditions. As the burden of multimorbidity increases, so does the individual’s likelihood of developing these outcomes. However, different measures of multimorbidity will exhibit varying degrees of association strength, thereby offering distinct predictive utility.

## Methods

### Participants

The MRC National Survey of Health and Development (NSHD) was established in 1946 and is an ongoing longitudinal population-based study, sampled from births recorded in England, Wales, and Scotland one week in March 1946 (from a total of 16,695 registered births).^33^ From these, a socially stratified sample of 5,362 (47.5% female) was selected at age 2 and followed since with data collections in adulthood at ages 26, 36, 43, 53 and 60-64 (63 on average) and 69 years.

### Chronic disease assessment

The clinical diagnoses of chronic diseases were derived from multiple sources of information of study members, involving self-reports, medication records, anthropometric data, and specific clinical measurements. Developing a clinically derived list of chronic diseases was fundamental in operationalizing multimorbidity. For this, the Delphi technique, recognised as a structured communication tool, was used to attain an informed consensus regarding the classification of conditions as chronic. The Delphi team comprised ten clinicians, each tasked with indicating their decision and providing it with a brief explanation using the same form (see Supplemental Table 1). At age 63, we derived definitions for: anaemia, cancer, coronary heart disease, diabetes, depression, dyslipidaemia, epilepsy, gastrointestinal disorders, hypertension, obesity, osteoarthritis, Parkinson’s disease, psychotic disorders, respiratory disorders, rheumatoid arthritis, and stroke. Conditions were coded as present/not present at age 36, 43, 53, and 63; For full description and definitions, see (Supplemental Table 1).

### Multimorbidity operationalizations

Various measures for multimorbidity operationalizations will be used to demonstrate the association to various outcomes (mortality and physical capability) and their relative predictive utility.

#### Binary multimorbidity

Based on multimorbidity defined as the co-occurrence of two or more diseases within an individual, we used the sum of reported conditions categorized into binary groups (0/1 and ≥2).

#### Unweighted disease count

This is the unweighted sum of the number of conditions out of the 16 specified, present in an individual.

#### Weighted disease count

Some studies have weighted each condition based on its burden before summing. Often weights are generated from within the analysis sample, which has drawbacks in terms of applicability to other datasets. We pooled weights across published studies,^34–36^ and indexes (i.e. Cambridge multimorbidity score, Charlson comorbidity score, Chronic disease score etc)^37–40^ using random-effect meta-analysis (see Supplemental Table 7), using this to define a weighted sum score.

#### Clustered multimorbidity

Using the International Classification of Disease 10^th^ Revision (ICD-10) system, we categorized the 9 index conditions into 7 physiological systems: *Cardiometabolic*: Diabetes, Hypertension, Obesity; *Neuropsychiatric*: Depression, Epilepsy; *Neoplasm*: Cancer; *Respiratory*: Respiratory disorders; *Digestive*: Gastrointestinal disorders; *Skin*: Dermatological disorders

Study members were classified into mutually exclusive groups:

a. two or more conditions affecting one body system (associative MM)
b. two or more conditions affecting two body systems (mixed MM)
c. three or more conditions affecting three or more body systems (complex MM), following the definition of complex multimorbidity used in various studies.^41,42^

#### Cumulative multimorbidity

To account for the duration of exposure to a given long-term condition, we made continuous variables for time since first incident report from each age (age 36, 43, 53, 63) up to age 63. We considered both linear and exponential weightings.

With linear weighting, weighted scores linearly increased with the increased total disease duration. It was assigned by dividing the total disease duration by 10. For example, if a study member had 20 years of diabetes and 10 years of hypertension, a score of 3 was given. Exponential weighting investigated if longer exposure to a long-term condition had geometrical impact on health. We applied the power of 1.1 to linear weighting. For example, using the same example above, the study member X would have a 3^1.1 = 3.35 score.

### Outcomes

#### Mortality

All-cause mortality data was obtained from all consenting study members linked to the National Health Service (NHS) central register, starting from the age of 26 and onward. The variables included their year of death, death in 3 months (e.g. January-March), and death status. For these analyses, follow-up time was from ages 60-64 years (dependent on the timing of the nurse visits or postal questionnaire) to mortality, or censored due to emigration or the end of September 2018.

#### Physical function

Physical function, which denotes an individual’s capacity to perform essential physical tasks necessary for daily functioning, is frequently evaluated through a combination of performance-based assessments and self-reported measures. We considered nurse-assessed personal and instrumental activities of daily living, and physical capability measures.

##### Functional independence

At age 60-64 (63 on average) and 69, we used the Office of Population Censuses and Surveys (OPCS) survey of disability, asking specifically about the difficulty with day-to-day activities. There were 20 items for general mobility, Activities of Daily Living (ADL) variables, and Instrumental Activities of Daily Living (IADL). These were summed to give a total disability score ranging from 0 (least disability) to 20 (worst disability) (see Supplemental Table 3).

##### Physical capability

We measured grip strength, chair rise time and standing balance during home visits at age 63 and 69 under the supervision of a trained nurse following standardized protocols.^43,44^ The selection of these three assessments was based on their widespread utilisation as fundamental indicators of physical capability within epidemiological research.^43^ These tests can detect meaningful variation in physical capability between individuals in late life across the full spectrum of ability (see Supplemental Table 6). A score was recorded for each performance-based measure if the study member demonstrated willingness and capability to undergo the specific test. When a study member was unwilling or unable to fulfil the task, the nurse documented the reason, subsequently moving on to the next task in the assessment sequence. These assessments were conducted at a regular pace, and study members were permitted to use personal aids if necessary; however, the use and type of aid were documented. Assistance from another person was not permitted.

### Covariates

Physical capability measures at 63 were used in the adjusted model to look at changes in physical capability from 63 to 69. Although sex and socioeconomic position were not included within the regression model, they were used in the descriptive study to look at stratified differences.

### Statistical analysis

We described the distribution and prevalence of multimorbidity using the operationalizations defined above. Predictive utility was estimated using survival analyses (outcome = mortality), zero-inflated Poisson regression (outcome = functional independence), and linear regression (outcome = physical capability).

#### Descriptive statistics

Descriptive analyses of the study population comprised the summary statistics to investigate overarching patterns. These were performed by generating histograms and frequency tables to evaluate the distribution, means, and range of each variable within the dataset. Each variable underwent examination for potential outliers, and normality was tested for continuous variables by histogram and Shapiro-Wilk test. The characteristics of study members were presented as absolute numbers and proportions (%), or as mean ± standard deviation (SD), or 95% confidence intervals (95% CI), as appropriate.

#### Survival analyses

From the dataset, two time variables were used; year at death and survival time (year at death minus year at completing the questionnaires either nurse or postal occurring between 2006-2010). Once the data were set as survival data, the longitudinal analysis approaches were used such as the Cox proportional hazards regression. The follow-up period for longitudinal analyses began between 2006-2010 to 2018: 8-12 years. In the analysis, the mortality rate was examined in relation to various multimorbidity measures at age 63. Cox proportional hazards modelling was used to test whether there were significant differences in rates between the sub-groups of the multimorbidity measures with the mortality. The proportional hazards assumption was checked amongst various multimorbidity measures graphically using Schoenfeld residuals.

#### Zero-inflated Poisson (ZIP) regression models

To test the effects of multimorbidity measures on independency; a count variable with excess zeros, and zero-inflated Poisson (ZIP) regression models were implemented for all multivariable models. In this study, the dependent variable was the count of disabilities observed among study members. Given the characteristics of the data on independence, the response of “no disability” was coded as 0 (zero count), and the probability of experiencing “no disability” represents the phenomenon of “zero-inflation.” Also, the models were fit for identical covariates (independency at 63) for both the count (number of disability scores) and inflate (no disability) portions of the ZIP model to assess the change in independency from age 63 to 69. Postestimation testing was undertaken to validate that the ZIP model stood as the most appropriate analytic approach when compared to linear, Poisson, and negative binomial regressions. The model coefficients were exponentiated to derive incidence rate ratios (IRRs), or the ratio of expected counts, which can be interpreted as a relative risk ratio. The models and estimates were interpreted based on p-values < 0.05.

#### Linear regression models

Linear regression analysis was used to assess the mean difference in physical performance associated with each multimorbidity measure. Before undertaking regression analysis, model assumptions were tested by examining diagnostic plots depicting the relationship between condition or multimorbidity and the continuous outcomes. The graphical analyses demonstrated that the relevant assumptions were satisfied.

The coefficients produced from linear regression where the exposure is categorical indicate the average difference in units of the outcome when moving from the reference category to any particular category. For each outcome, the corresponding regression model was conducted with multimorbidity measures as exposure variables. Analyses were initially conducted unadjusted and then included age 63 physical limitation for changes in physical functioning in the adjusted model

## Results

### Descriptives

The sample included 2,653 individuals (51.6% female) (Table 1). 1,702 (64.2%) had *binary multimorbidity* with a similar prevalence in both sexes. Only 353 (13.3%) individuals did not have any condition. Three clusters were identified: 248 (9.4%) individuals had associative multimorbidity, 806 (30.4%) had mixed multimorbidity, and 648 (24.4%) had complex multimorbidity.

**Table 1:**
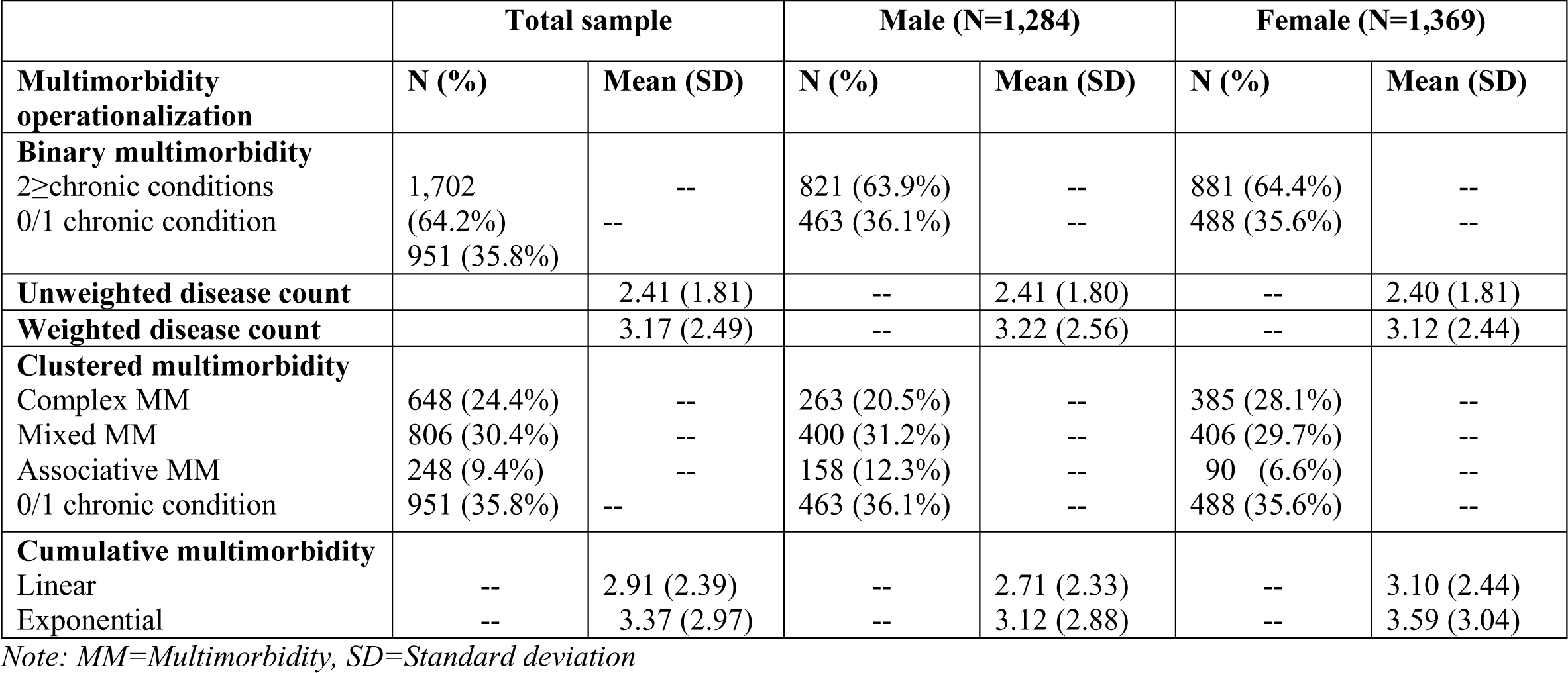
Descriptive statistics of different multimorbidity operationalizations at age 63, n=2653.

### Multimorbidity and mortality

There were 172 deaths (6.48%) over a median 4.7 years of follow-up, giving a 5.2/1000 person-years mortality rate (Table 2). Study members with basic multimorbidity had a lower survival probability than those without (HR 1.76 (95% CI 1.24 to 2.49)). Higher *unweighted disease count* was associated with higher mortality, either continuous (HR 1.16 per disease (1.07 to 1.24)) or weighted (HR 1.12 (1.06 to 1.18)). Complex multimorbidity was associated with the highest mortality (HR 1.85 (1.23 to 2.78)). The HR for mortality with cumulative multimorbidity per one unit increase was 1.06 (1.00 to 1.13) for a linear and 1.05 (1.00 to 1.10) for an exponentiated model.

**Table 2:** The relationship between multimorbidity measures in predicting mortality.

| **Multimorbidity operationalization** | **Categories** | **Number of deaths** | **HR (95% CI)** | **P-value** |
| --- | --- | --- | --- | --- |
| **Binary multimorbidity** | 0/1 N=951 | 42 (4.4%) | 1.00 (Ref.) | -- |
|  | 2≥ N=1,702 | 130 (7.6%) | 1.76 (1.24 to 2.49) | **0.002** |
| **Unweighted disease count** | N=2653 | 172 (6.5%) | 1.15 (1.07 to 1.24) | **0.001** |
| **Weighted disease count** | N=2653 | 172 (6.5%) | 1.12 (1.06 to 1.18) | **0.001** |
| **Clustered multimorbidity** | 0/1 N=951 | 42 (4.4%) | 1.00 (Ref.) | -- |
|  | AssociativeMM N=248 | 20 (8.1%) | 1.84 (1.08 to 3.13) | **0.025** |
|  | MixedMM N=806 | 58 (7.2%) | 1.65 (1.11 to 2.46) | **0.013** |
|  | ComplexMM N=648 | 52 (8.0%) | 1.85 (1.23 to 2.78) | **0.024** |
| **Cumulative multimorbidity** | Linear N=2653 | 172 (6.5%) | 1.06 (1.00 to 1.13) | **0.038** |
|  | Exponential N=2653 | 172 (6.5%) | 1.05 (1.00 to 1.10) | **0.033** |

*Note: MM= Multimorbidity*

**Table 3:**
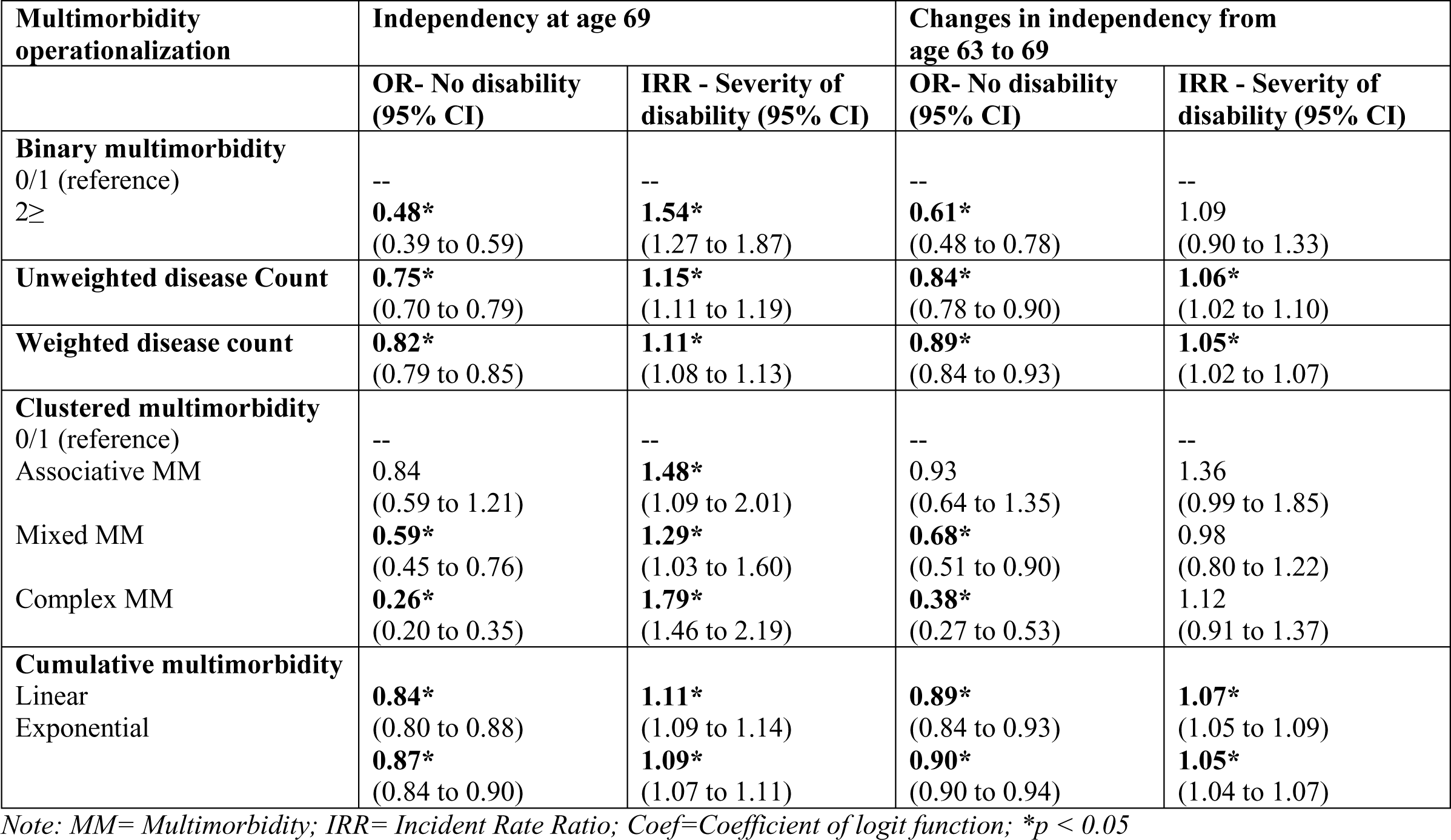
Zero-inflated Poisson regression coefficients and confidence interval, for the odds of no disability (inflate) and the disability scores (count), at age 69, and changes from age 63-69.

**Table 4:** Linear regression coefficients (unstandardized) and confidence interval for multimorbidity measures and physical performances, at age 69, and changes from age 63-69.

| Multimorbidity measures | Grip strength @ 69 | ΔGrip strength @ 63-69 | Chair rise speed @69 | Δ Chair rise speed @63-69 | Standing balance @69 | Δ Standing balance @63-69 |
| --- | --- | --- | --- | --- | --- | --- |
| **Binary multimorbidity**<br>0/1 (reference)<br>2≥ | --<br>**-2.59\***<br>(-3.54 to -1.65) | --<br>**-1.28\***<br>(-1.95 to -0.60) | --<br>**-0.04\***<br>(-0.06 to -0.02) | --<br>**-0.04\***<br>(-0.06 to 0.01) | --<br>**-0.65\***<br>(-0.98 to -0.32) | --<br>**-0.59\***<br>(-0.94 to -0.25) |
| **Unweighted disease Count** | **-0.80\***<br>(-1.06 to -0.54) | **-0.45\***<br>(-0.63 to -0.27) | **-0.02\***<br>(-0.03 to -0.01) | **-0.01\***<br>(-0.02 to 0) | **-0.27\***<br>(-0.36 to -0.18) | **-0.24\***<br>(-0.34 to -0.14) |
| **Weighted disease count** | **-0.55\***<br>(-0.74 to -0.37) | **-0.32\***<br>(-0.46 to -0.19) | **-0.01\***<br>(-0.02 to 0) | **-0.01\***<br>(-0.02 to 0) | **-0.15\***<br>(-0.22 to -0.08) | **-0.14\***<br>(-0.21 to -0.07) |
| **Clustered multimorbidity**<br>0/1 (reference)<br>Associative MM<br>Mixed MM<br>Complex MM | --<br>0.43<br>(-1.27 to 2.13)<br>**-2.04\***<br>(-3.16 to -0.92)<br>**-4.48\***<br>(-5.69 to -3.28) | --<br>-0.20<br>(-1.39 to 0.99)<br>-0.82<br>(-1.60 to -0.04)<br>**-2.27\***<br>(-3.11 to -1.43) | --<br>**-0.04\***<br>(-0.08 to -0.01)<br>-0.02<br>(-0.05 to 0.01)<br>**-0.06\***<br>(-0.09 to -0.04) | --<br>**-0.05\***<br>(-0.09 to -0.01)<br>**-0.03\***<br>(-0.05 to -0.01)<br>**-0.04\***<br>(-0.07 to -0.02) | --<br>**-0.84\***<br>(-1.44 to -0.25)<br>-0.33<br>(-0.72 to 0.07)<br>**-1.00\***<br>(-1.43 to -0.57) | --<br>**-0.84\***<br>(-1.45 to -0.23)<br>-0.30<br>(-0.70 to 0.10)<br>**-0.89\***<br>(-1.32 to -0.45) |
| **Cumulative multimorbidity**<br>Linear<br>Exponential | **-0.76\***<br>(-0.95 to -0.56)<br>**-0.62\***<br>(-0.77 to -0.46) | **-0.39\***<br>(-0.52 to -0.25)<br>**-0.31\***<br>(-0.43 to -0.20) | **-0.01\***<br>(-0.02 to 0)<br>**-0.01\***<br>(-0.02 to 0) | **-0.01\***<br>(-0.02 to 0)<br>**-0.01\***<br>(-0.02 to 0) | **-0.20\***<br>(-0.27 to -0.13)<br>**-0.16\***<br>(-0.21 to -0.10) | **-0.16\***<br>(-0.23 to -0.09)<br>**-0.13\***<br>(-0.19 to -0.07) |

Note: MM= Multimorbidity; \* $p < 0.05$ ;

Somers’ D (Figure 1), similar to R2, gives explained variation to compare which model has the highest predictive power in Cox regression. *Unweighted disease count* (16.3%) and *weighted disease count* (16.7%) have shown the highest predictive power for mortality compared to other operationalizations.

**Figure 1:**
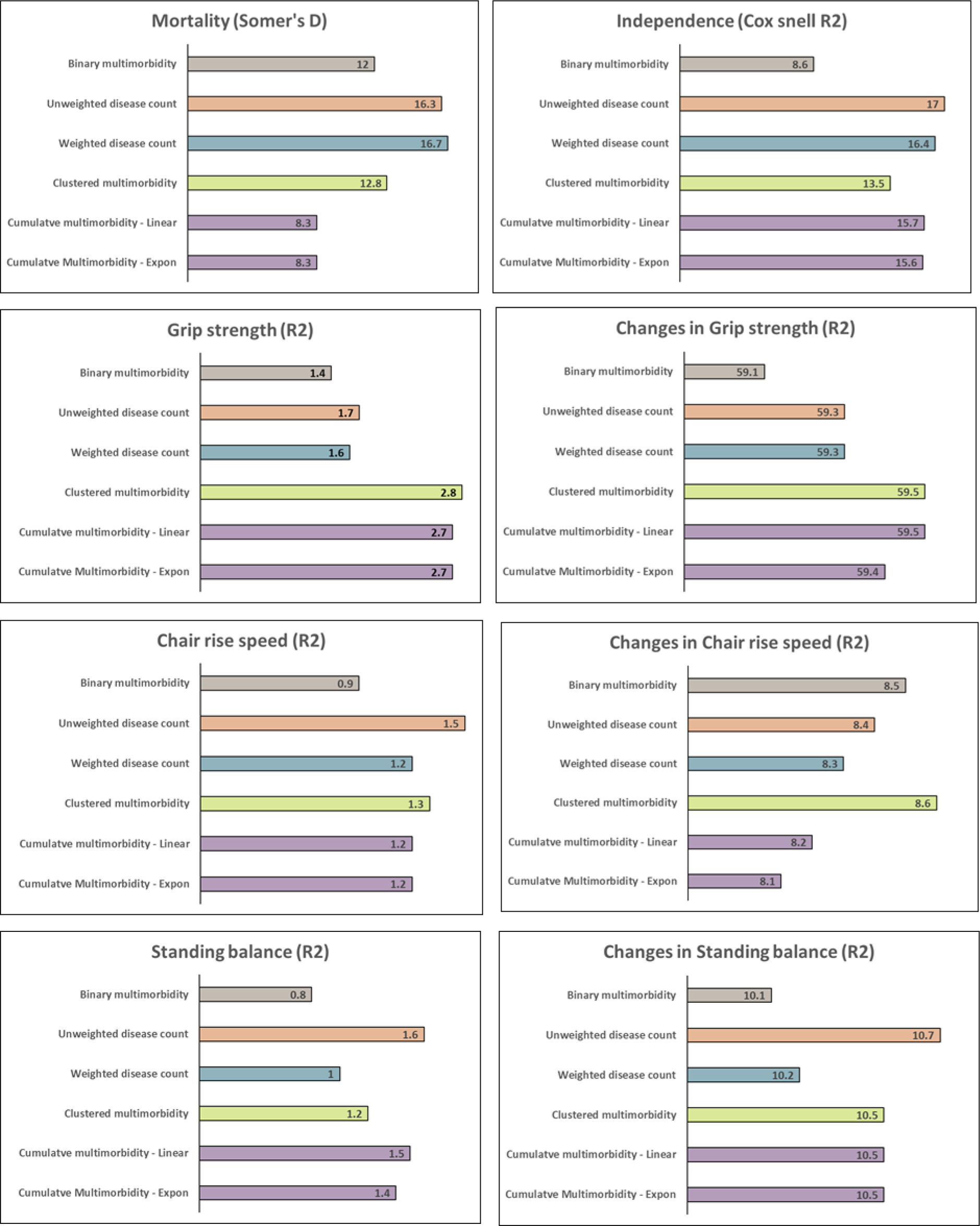
Explained variation for which multimorbidity measures has the highest predictive utility for mortality (Somer’s D), independency (ML cox-snell), and physical performances (R-squared).

### Multimorbidity and physical function

Across the 20 measures of functional independence at age 69, 46.5% (N=999) of the study members at age 69 reported no difficulty, and 24.4% (N=525) reported one difficulty in all measures. A graded pattern was observed among those who reported difficulty for each task, with fewer people reporting more problems. These results suggest that there may be some evidence of a ceiling effect in the self-reported measures, with the majority of study members reporting no difficulty with the 20 tasks assessed. The mean functional independence score was 1.66 and there was a presence of excess zero counts (46.5%).

In the analysis of the *unweighted disease count, weighted disease count, and cumulative multimorbidity* showed significant associations to both functional independence at age 63 and changes in functional independence. Higher scores on multimorbidity measures were generally associated with increased incidence of difficulties in functional independence tasks and increased changes in functional independence. The results of the logit model analysis indicated that no-disability group had lower scores on *unweighted disease count, weighted disease count,* and *cumulative multimorbidity*. In general, lower scores on multimorbidity measures were associated with an increased possibility of not having difficulties in independency tasks and not having changes in independency.

Cox-Snell R2 (Figure 1), gives explained variation to compare which model has the highest predictive power for zero-inflated Poisson regression. *Unweighted disease count* (17.0%) and *weighted disease count* (16.4%), similar to Somer’s D in Cox regression, have shown the highest predictive power for functional independence than any other measures within the exposure variables.

In the linear regression model analysis, *unweighted disease count, weighted disease count,* and *cumulative multimorbidity* showed significant associations to both physical capabilities and changes in physical capabilities. Higher scores on multimorbidity measures were generally associated with poorer physical capabilities and longitudinal decrease in physical capabilities.

## Discussion and Implications

This study examined various multimorbidity operationalizations to compare predictive utilities for mortality and physical functioning in a longitudinal sample. Different multimorbidity measures, in general, were all associated with higher mortality and poorer physical function. *Unweighted disease count* demonstrated the highest predictive utility or variation explained. Both *weighted* and *cumulative multimorbidity* did not add any predictive value over the *unweighted disease count*. *Clustered multimorbidity* was less predictive of mortality outcomes but better predicted changes in physical functioning markers such as grip strength. Although most commonly used in multimorbidity studies, *binary multimorbidity* was insufficiently useful overall.

The higher mortality risk among study members with increasing disease count found in our study is consistent with other data showing a dose-response relationship.^7^

Previous studies have demonstrated that multimorbidity is linked to functional limitations both cross-sectionally and longitudinally.^26–32^ We found that various operationalizations of multimorbidity were all associated with lower functional independence and physical capability. Moreover, individuals with more conditions, more severe diseases and specific disease patterns experienced larger changes in functional decline.^8^ Together, these contrast with research reporting no association between comorbidity^47^ or grouped multimorbidity^48^ with physical functioning. One similar cohort study showed at least 50% of the participants were functionally independent despite chronic disorders among older people over 65.^49^

Our data simultaneously compare different strengths of association between multimorbidity definitions and mortality, functional independence and physical capability. Quantifying how these vary offers practical choices between methods of operationalizing multimorbidity for various purposes in research and clinical settings, for example a targeted strategy for understanding mortality, physical function and health care utilization for older people.

Our analyses benefit from prospective ascertainment of multimorbidity and serial measures of functional independence. We included many common chronic conditions, using the same conditions across different operationalizations. However, we were sometimes limited by questionnaire phrasing, where disease were often aggregated (e.g. respiratory disorders or gastrointestinal disorders), which could obscure relationships. The limited number of included conditions reinforces the need to corroborate these findings in much larger datasets with a larger number of conditions, such as electronic health records.

There have been changes in disease definitions and criteria over time, which makes it difficult to study some conditions in greater detail. For example, we had to group all respiratory conditions to account for different measurement methods over the last thirty years. Some conditions like coronary heart disease, stroke and kidney disorders were defined by self-report. However, self-reported diagnoses do not always lead to bias, and we have previously found >90% agreement for diabetes diagnosis between self-report and primary care diagnosis in this cohort.^50^ In future, we may overcome general concerns over biased ascertainment by triangulating multiple sources, including self-report, electronic health records and biomarker assessments at multiple time points.

Multimorbidity research urgently needs a standardized approach to its measurement and operationalization. We add to this possibility by using predictive utility for mortality, functional independence and physical capability as criteria for determining the usefulness of a given operationalization. We showed that *unweighted disease count* was best in this respect, with the advantage of being conceptually straightforward and more likely to be robust.

## Funding

This work was supported by the following grants: This work was supported by the UK Medical Research Council which provides core funding for the Medical Research Council Unit for Lifelong Health and Ageing at University College London and the Medical Research Council National Survey of Health and Development (grant numbers MC_UU_00019/1, MC_UU_00019/3). Han Sang Seo was supported by grant from Medical Research Council (grant number MC_UU_00019/3).

## Acknowledgements

We are grateful to the cohort members in the 1946 National Study of Health and Development who have provided data for research throughout their lives for their continuing support. We also thank members of the NSHD scientific and data collection teams.

## Conflict of Interest

None declared.

## Data Availability

The 1946 MRC National Survey of Health & Development (NSHD) data is available for research via application to the NSHD Data Sharing Committee, a data sharing agreement being in place between UCL and the academic institution that employs the researcher, and LHA resources being available to meet the requests for data sharing. Additionally, all proposals to use the NSHD data must support and adhere to the core principles of data sharing with the Medical Research Council (MRC).

For more information on the NSHD, access to data & collaborations, please see http://www.nshd.mrc.ac.uk/data/data-sharing or contact

Further information can also be accessed at: https://skylark.ucl.ac.uk

## Author Contributions

All authors conceptualized the research aims and contributed to the design of the study. HS prepared the dataset and conducted the analysis. HS and PP drafted the paper with input from all authors. All authors discussed the findings, contributed to drafting and read and approved the final manuscript.

## Supplementary file

**Supplemental Table 1.**
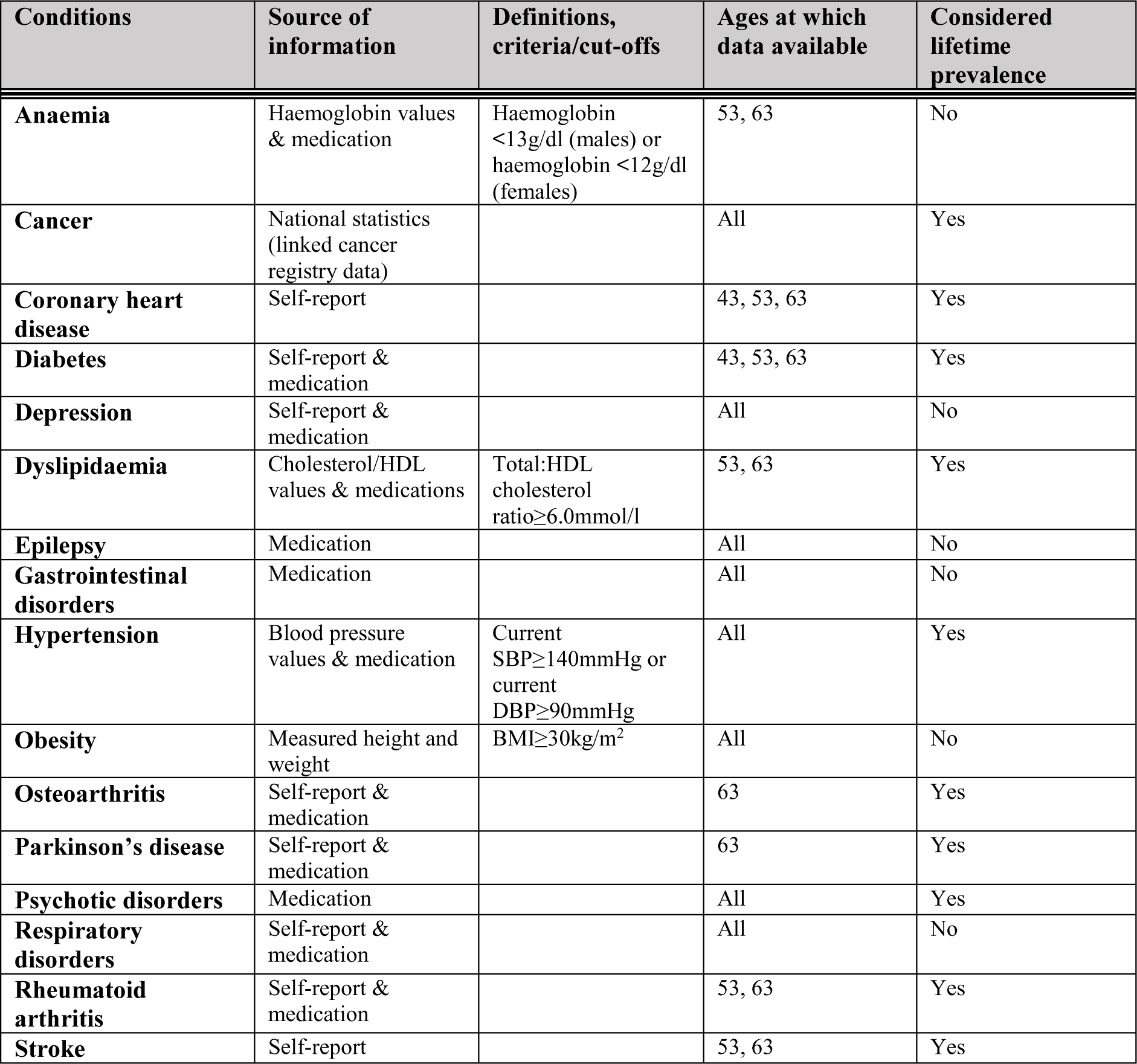
16 conditions contributing to multimorbidity score, with data sources and definitions.

| Conditions | Source of information | Definitions, criteria/cut-offs | Ages at which data available | Considered lifetime prevalence |
| --- | --- | --- | --- | --- |
| **Anaemia** | Haemoglobin values & medication | Haemoglobin <13g/dl (males) or haemoglobin <12g/dl (females) | 53, 63 | No |
| **Cancer** | National statistics (linked cancer registry data) |  | All | Yes |
| **Coronary heart disease** | Self-report |  | 43, 53, 63 | Yes |
| **Diabetes** | Self-report & medication |  | 43, 53, 63 | Yes |
| **Depression** | Self-report & medication |  | All | No |
| **Dyslipidaemia** | Cholesterol/HDL values & medications | Total:HDL cholesterol ratio≥6.0mmol/l | 53, 63 | Yes |
| **Epilepsy** | Medication |  | All | No |
| **Gastrointestinal disorders** | Medication |  | All | No |
| **Hypertension** | Blood pressure values & medication | Current SBP≥140mmHg or current DBP≥90mmHg | All | Yes |
| **Obesity** | Measured height and weight | BMI≥30kg/m<sup>2</sup> | All | No |
| **Osteoarthritis** | Self-report & medication |  | 63 | Yes |
| **Parkinson's disease** | Self-report & medication |  | 63 | Yes |
| **Psychotic disorders** | Medication |  | All | Yes |
| **Respiratory disorders** | Self-report & medication |  | All | No |
| **Rheumatoid arthritis** | Self-report & medication |  | 53, 63 | Yes |
| **Stroke** | Self-report |  | 53, 63 | Yes |

**Supplemental Figure 1.**
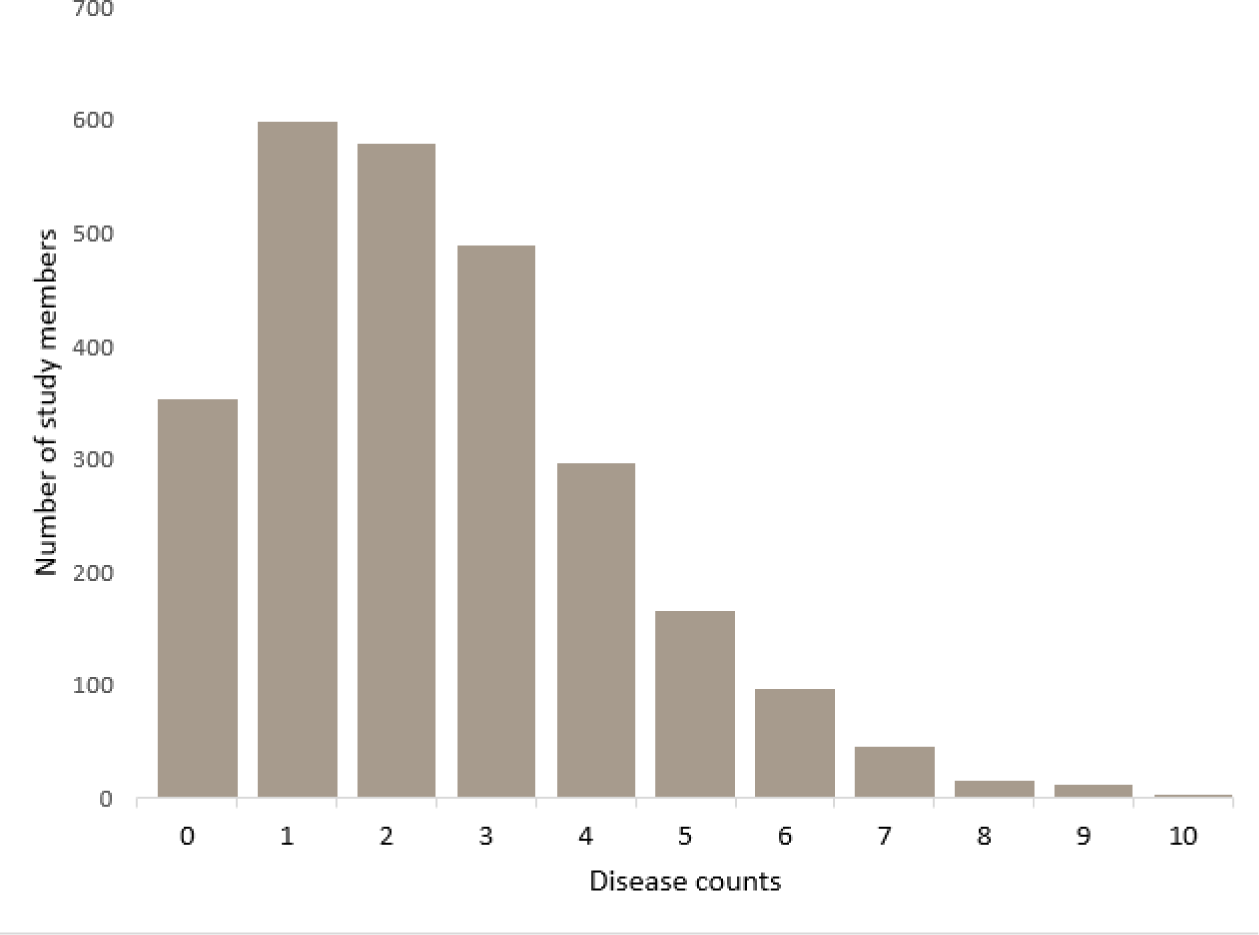
Distribution of disease count at age 63.

**Supplemental Table 2:**
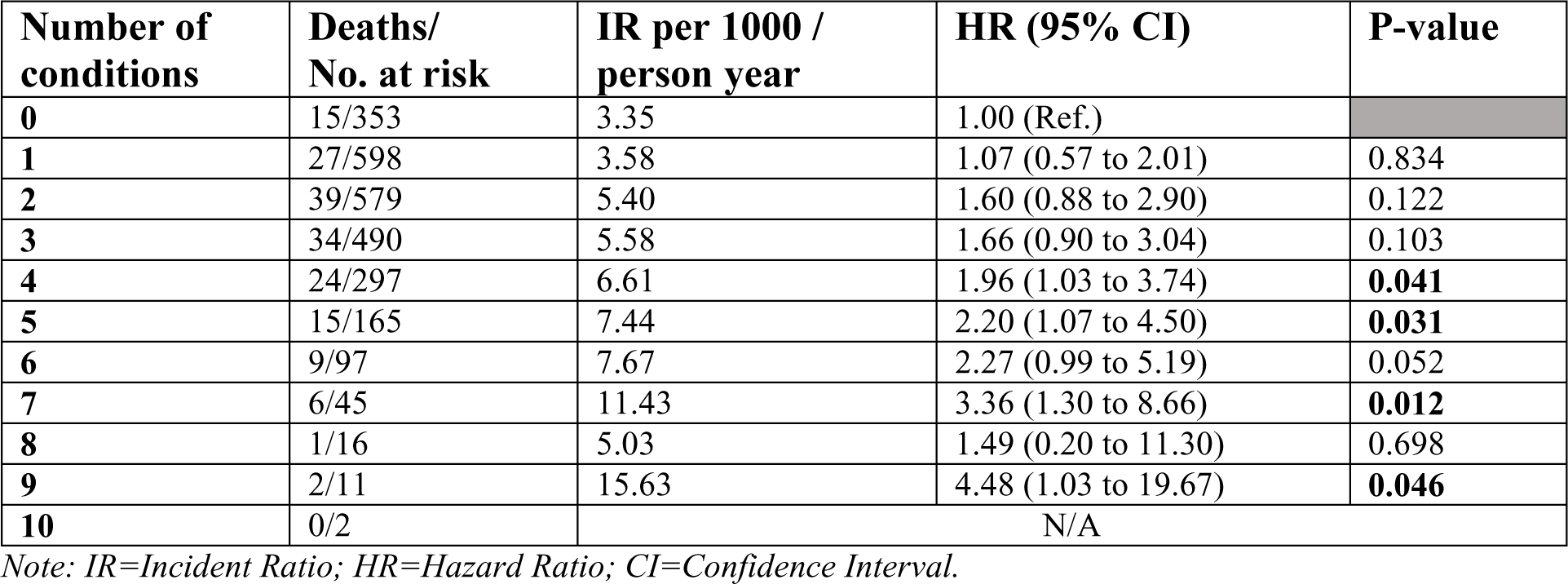
Association between disease count (category) and 12-year mortality.

**Supplemental Table 3.** 20 task lists included in the independency assessment at age 69.

| **Independency** |  |
| --- | --- |
| Walk ¼ mile | Shopping/carry bag |
| Climb 12 flight of stairs | House work, laundry |
| Arms to reach/stretch | Prepare meal |
| Holding, Grip, turning | Feeding |
| Bending down | Washing face and hand |
| Keeping balance | Dressing |
| Paperwork, bills | Cut toe |
| Getting out house | Getting in/out chair |
| Taking medicine | Getting in/out bed |
| Getting around indoor | Bathing |

### Physical capability

Several physical capability measures were assessed during home visits at age 63 and 69. Under the supervision of a trained nurse, and following standardised protocols, three performance-based measures were collected at both time points: grip strength, chair rise time and standing balance. These three tests were chosen for assessment because they are some of the most commonly used measures of physical capability in epidemiological studies and were expected to detect meaningful variation in capability between individuals in late-life across the full spectrum of ability.

For each physical capability measures a score was recorded if the study member was willing and able to perform that particular test. If the study member was unable or unwilling to complete the task the reason was noted and the nurse progressed onto the next task.

To measure grip strength, study members were asked to squeeze the handle of a Nottingham electronic handgrip dynamometer: three tests in their dominant hand and three in their non-dominant hand. The dynamometers are accurate, linear and stable to ±0.5kg, with each machine calibrated at the start of the assessment using a back-loading rig. The retest variability within individual study members for maximal voluntary tests of strength in those unused to such measurements is about 9%. The nurse provided strong vocal encouragement throughout the test to elicit maximal performance. The protocol differed slightly between data collection rounds. Two values were recorded for each hand and the highest used in analyses.

To measure chair rise time, study members were asked to perform 10 chair rises with their arms folded, as quickly as possible, and the time taken to complete the task, was recorded in seconds. The test was conducted using an armless, straight backed hard chair, with the seat approximately 46cm above the floor. Chair rise time was measured with a stopwatch as the time taken to rise from a sitting to a standing position with straight back and legs and then to sit down again 10 complete times as fast as possible. To ensure that all performance-based variables used followed a similar scale, with low values representing poor functional performance, the time taken to complete the task was converted into a measure of speed (number of rises per second).

To measure standing balance, study members were asked to fold their arms, stand on their preferred leg and raise their other leg a few inches above the ground, holding this position for a maximum of 30 seconds. The task was then repeated with the study members closing their eyes. For each test, the maximum time, up to 30 seconds, that the study member maintained the balance position was recorded. In line with previous work on these measures, performance time from the eyes-closed balance test were used, as a substantial ceiling effect was noted for the eyes-open test.

These tasks were performed at normal pace, and study members were allowed the use of personal aids if required, although the use and type of aid were noted. Help from another individual was prohibited.

**Supplemental Table 4a:** Sample characteristics of independency at age 69, n=2149.

|  | **All (N=2,149)** | **Male (N=1,052)** | **Female (N=1,097)** | **Sex differences** |  |
| --- | --- | --- | --- | --- | --- |
|  | **Mean (SD)** | **Mean (SD)** | **Mean (SD)** | **t** | **P-value** |
| **Independency** | 1.66 (2.81) | 1.28 (2.67) | 2.03 (2.90) | t = -6.23 | **<0.001** |

**Supplemental Table 4b:**
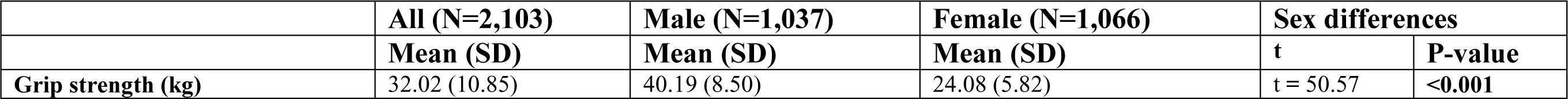
Sample characteristics of grip strength at age 69, n=2103.

**Supplemental Table 4c:** Sample characteristics of chair rise speed at age 69, n=1905.

|  | **All (N=1,905)** | **Male (N=926)** | **Female (N=979)** | **Sex differences** |  |
| --- | --- | --- | --- | --- | --- |
|  | **Mean (SD)** | **Mean (SD)** | **Mean (SD)** | **t** | **P-value** |
| **Chair rise speed (rise/s)** | 0.46 (0.22) | 0.46 (0.14) | 0.45 (0.27) | t = 0.77 | 0.44 |

**Supplemental Table 4d:**
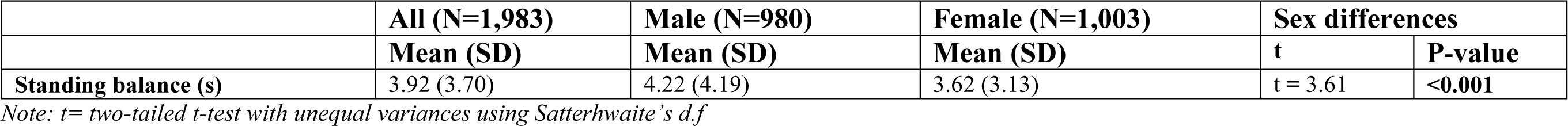
Sample characteristics of standing balance (eye closed), at age 69, n=1983.

|  | **All (N=1,983)** | **Male (N=980)** | **Female (N=1,003)** | **Sex differences** |  |
| --- | --- | --- | --- | --- | --- |
|  | **Mean (SD)** | **Mean (SD)** | **Mean (SD)** | **t** | **P-value** |
| **Standing balance (s)** | 3.92 (3.70) | 4.22 (4.19) | 3.62 (3.13) | t = 3.61 | **<0.001** |

*Note: t= two-tailed t-test with unequal variances using Satterhwaite's d.f*

**Supplemental Table 5.** Explained variations for which multimorbidity measures has the highest predictive utility for mortality (Somers’ D) and independency (ML cox-snell)

| Multimorbidity measures | Somer's D | ML(Cox-Snell) R2 |
| --- | --- | --- |
| Binary multimorbidity | 0.120 | 0.086 |
| Unweighted disease Count | 0.163 | 0.170 |
| Weighted disease count | 0.167 | 0.164 |
| Clustered multimorbidity | 0.128 | 0.135 |
| Cumulative multimorbidity |  |  |
| Linear | 0.083 | 0.157 |
| Exponential | 0.083 | 0.156 |

*Note: MM= Multimorbidity*

**Supplemental Table 6.** Explained variations for which multimorbidity measures has the highest predictive utility for physical performances (R-squared)

| **Multimorbidity measures** | **Grip strength @ 69** | **Δ Grip strength @ 63-69** | **Chair rise speed @69** | **Δ Chair rise speed @63-69** | **Standing balance @69** | **Δ Standing balance @63-69** |
| --- | --- | --- | --- | --- | --- | --- |
| **Binary multimorbidity** | 0.014 | 0.591 | 0.009 | 0.085 | 0.008 | 0.101 |
| **Unweighted disease Count** | 0.017 | 0.593 | 0.015 | 0.084 | 0.016 | 0.107 |
| **Weighted disease count** | 0.016 | 0.593 | 0.012 | 0.083 | 0.010 | 0.102 |
| **Clustered multimorbidity** | 0.028 | 0.595 | 0.013 | 0.086 | 0.012 | 0.105 |
| **Cumulative multimorbidity** |  |  |  |  |  |  |
| Linear | 0.027 | 0.595 | 0.012 | 0.082 | 0.015 | 0.105 |
| Exponential | 0.027 | 0.594 | 0.012 | 0.081 | 0.014 | 0.105 |

**Supplemental Table 7.**
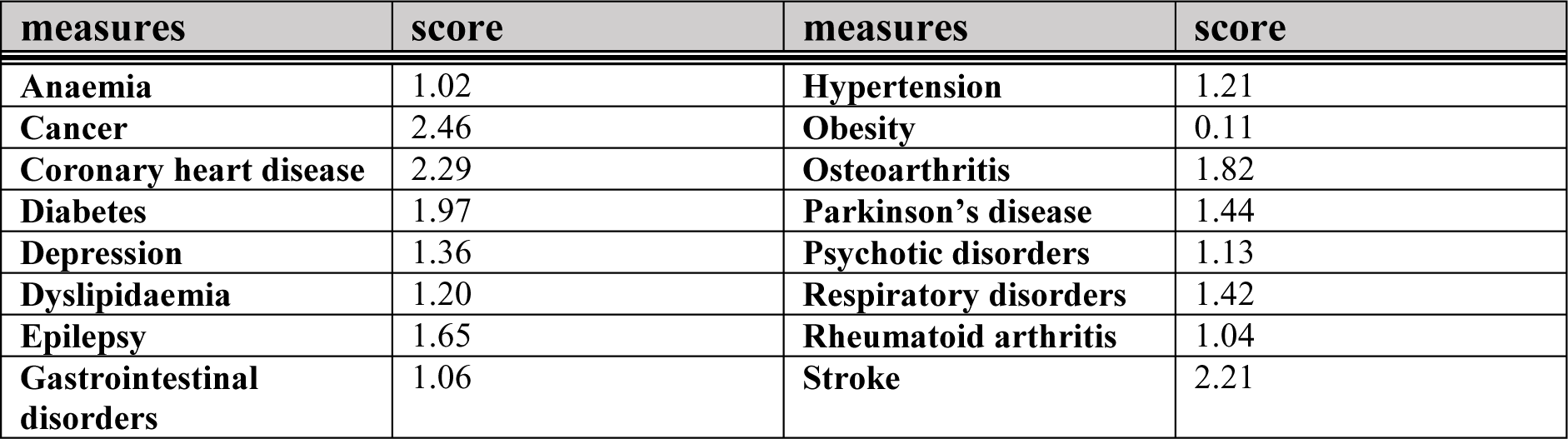
Pooled weighted multimorbidity score at age 63.

**Supplemental Figure 2.**
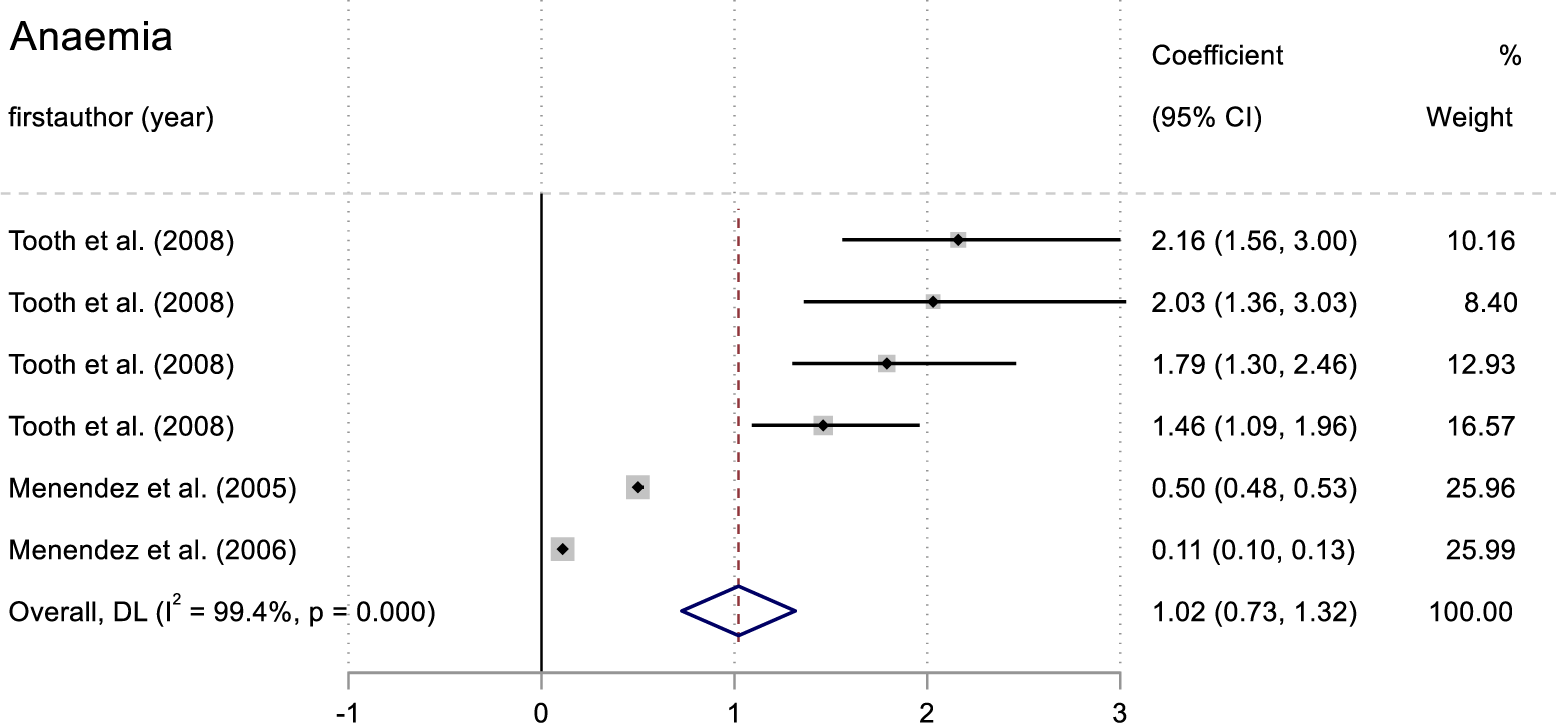
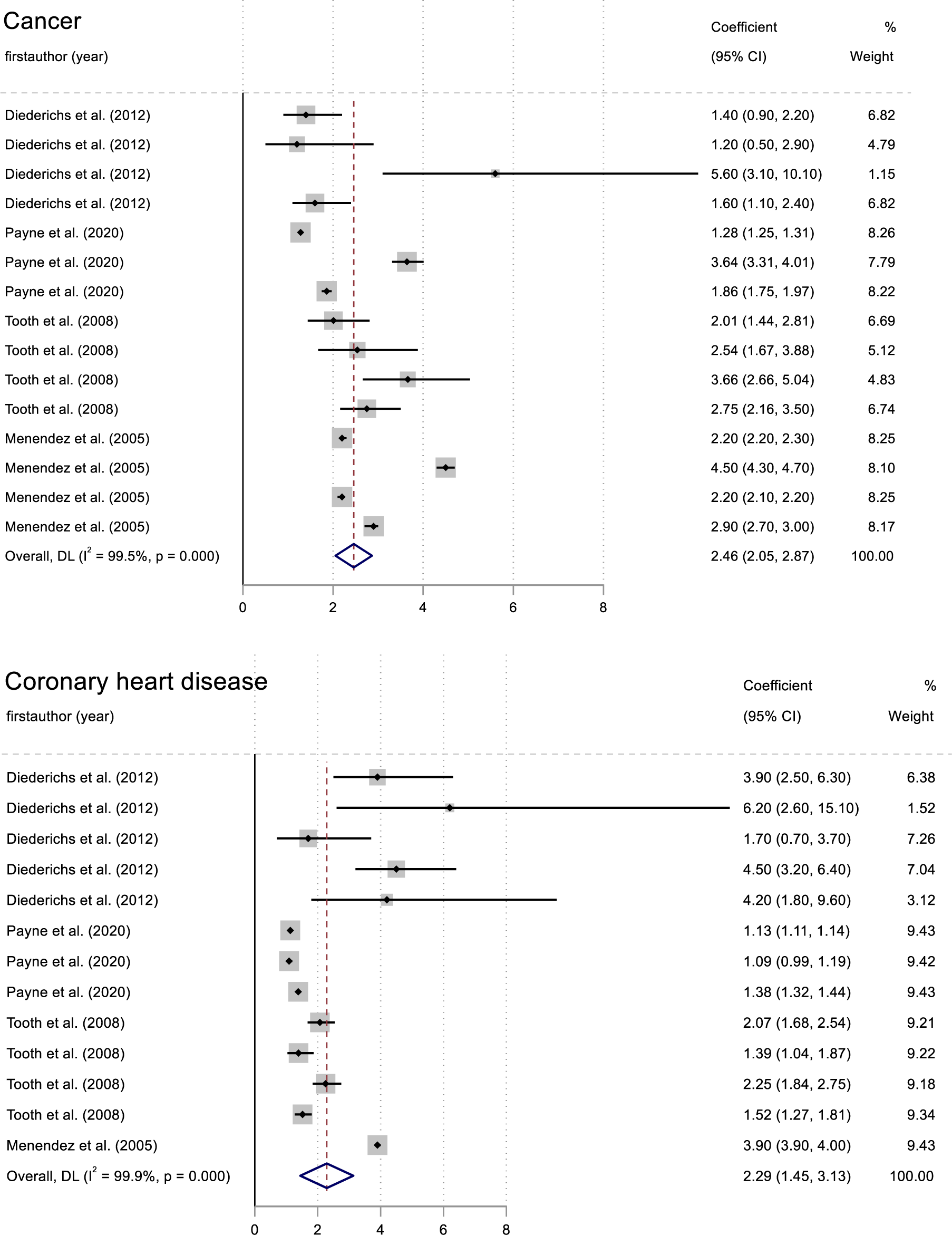
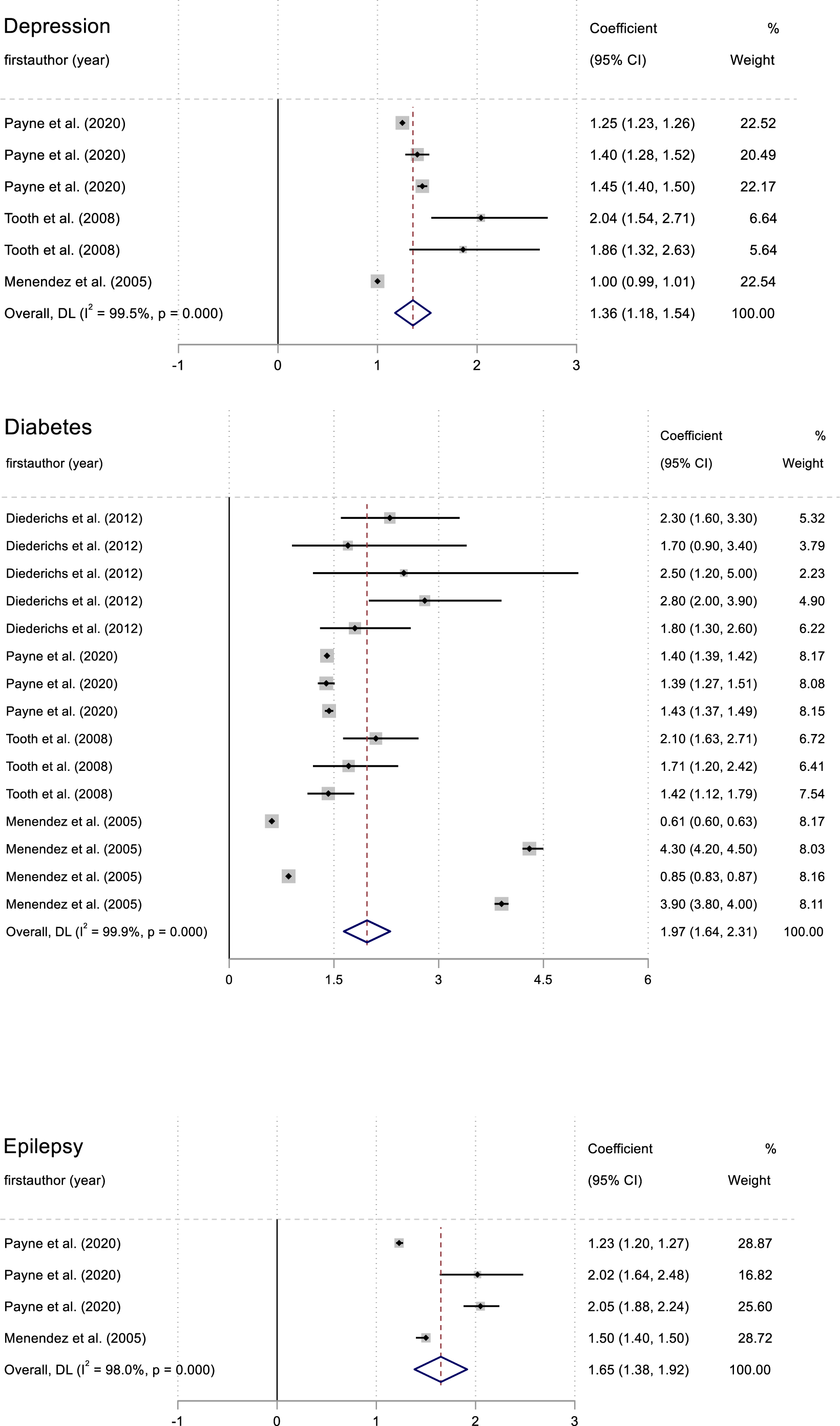
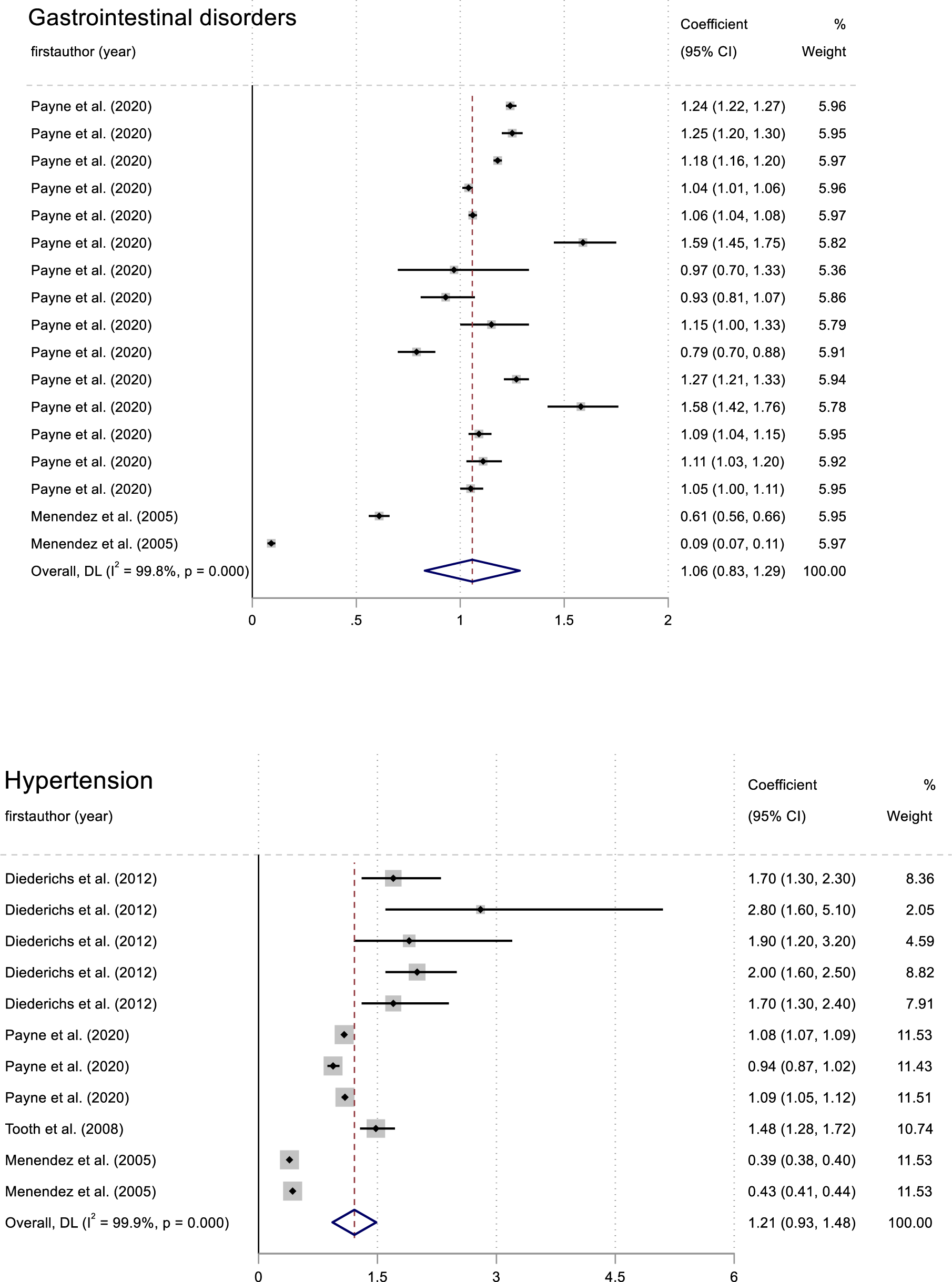
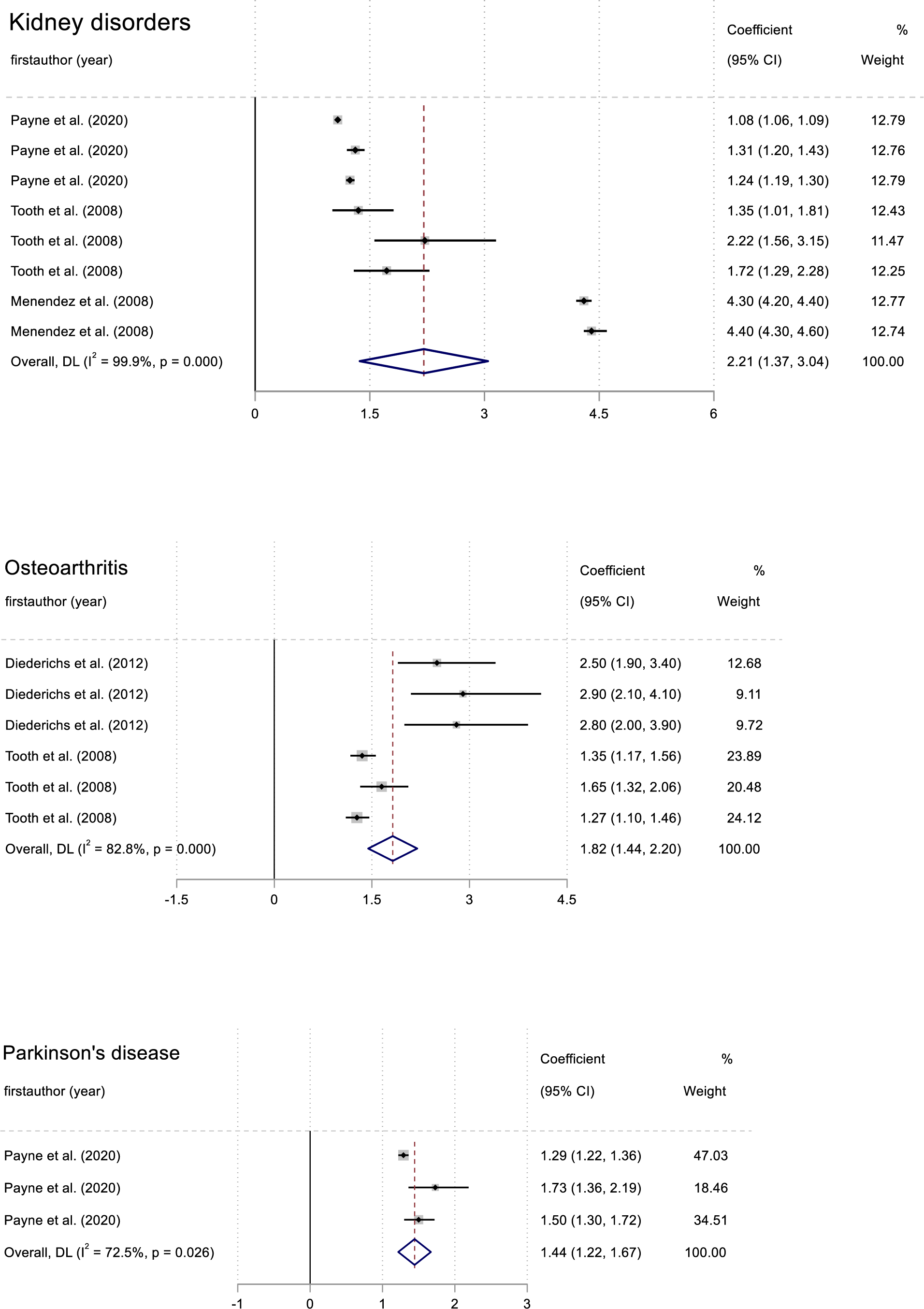
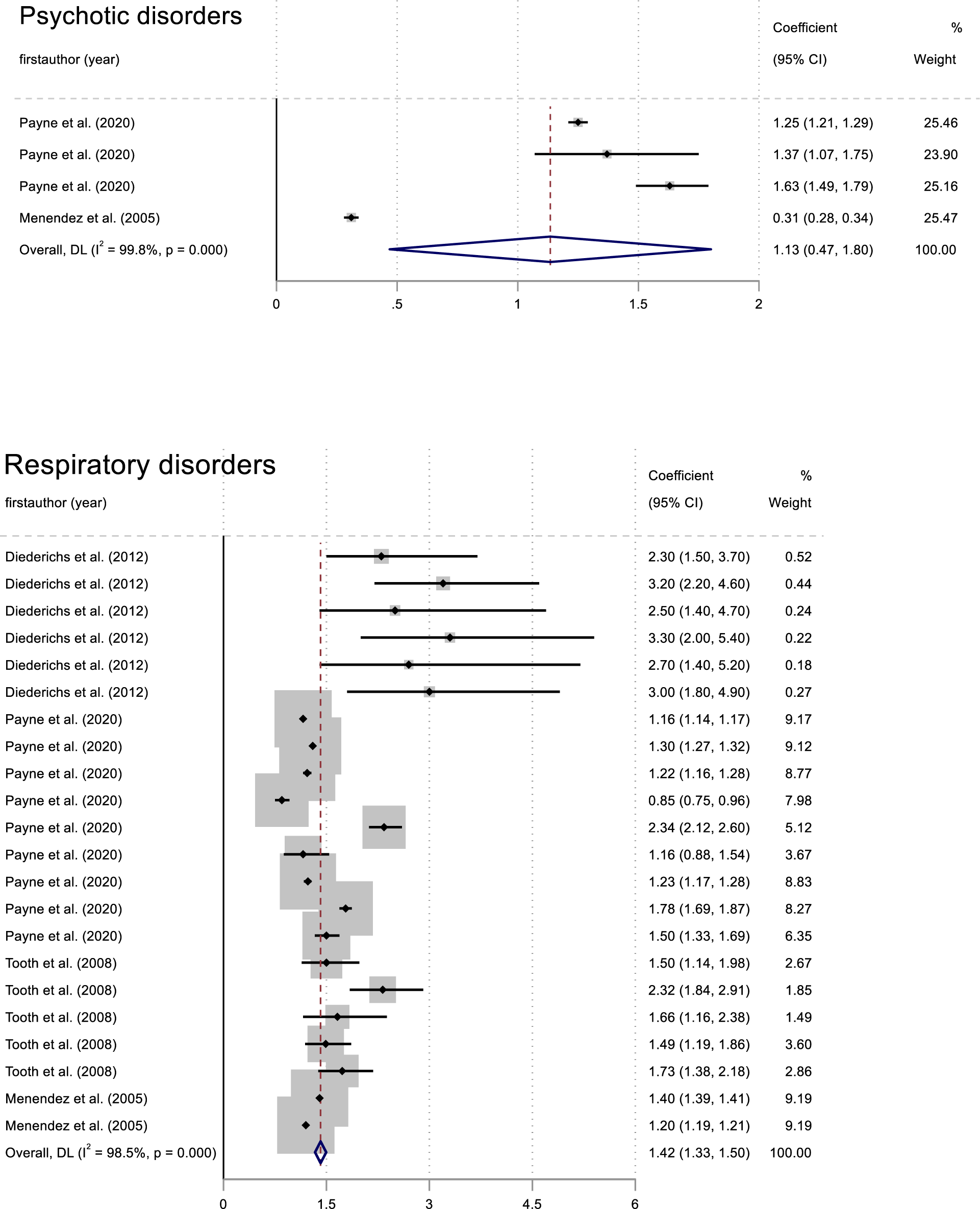
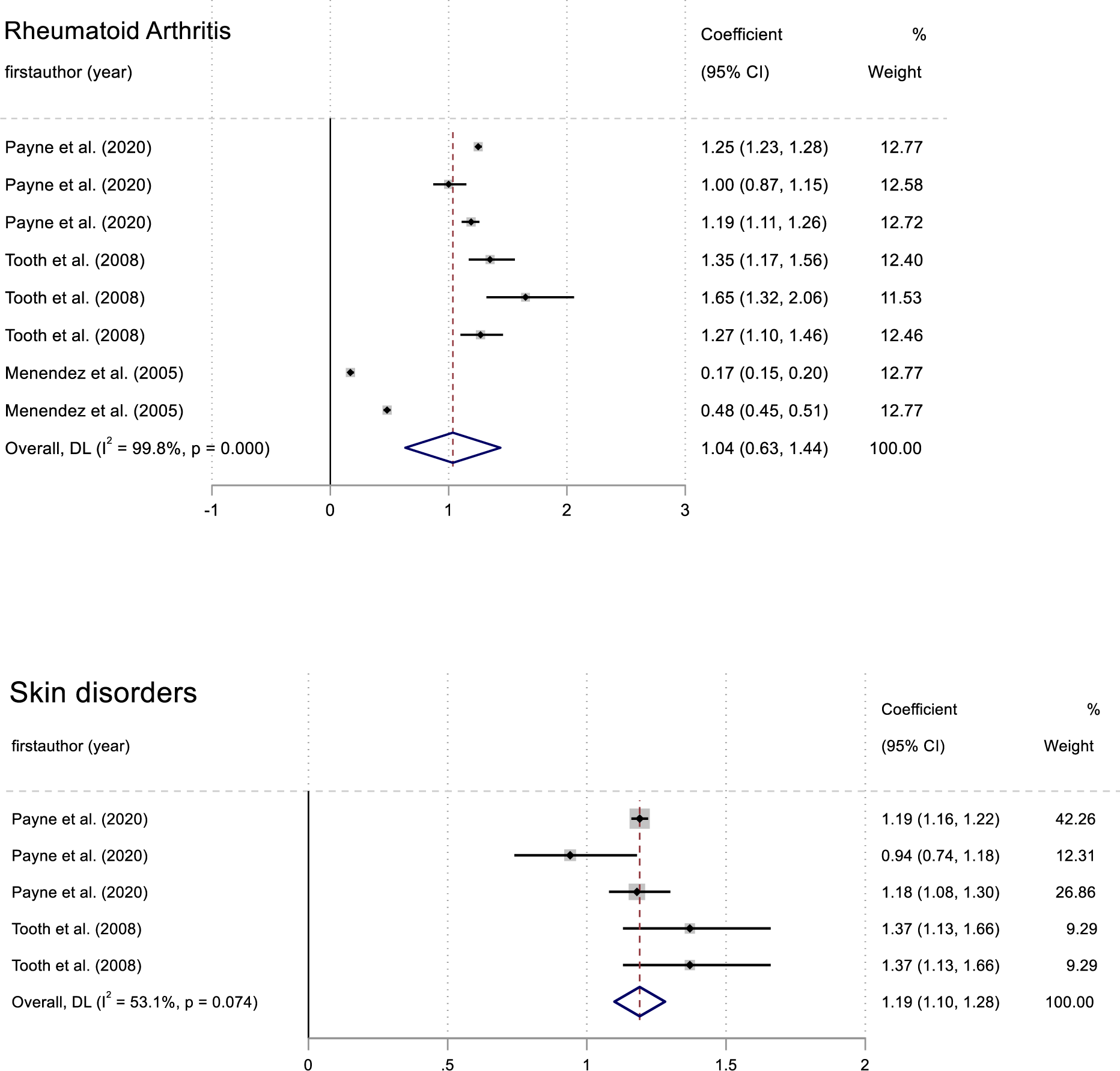
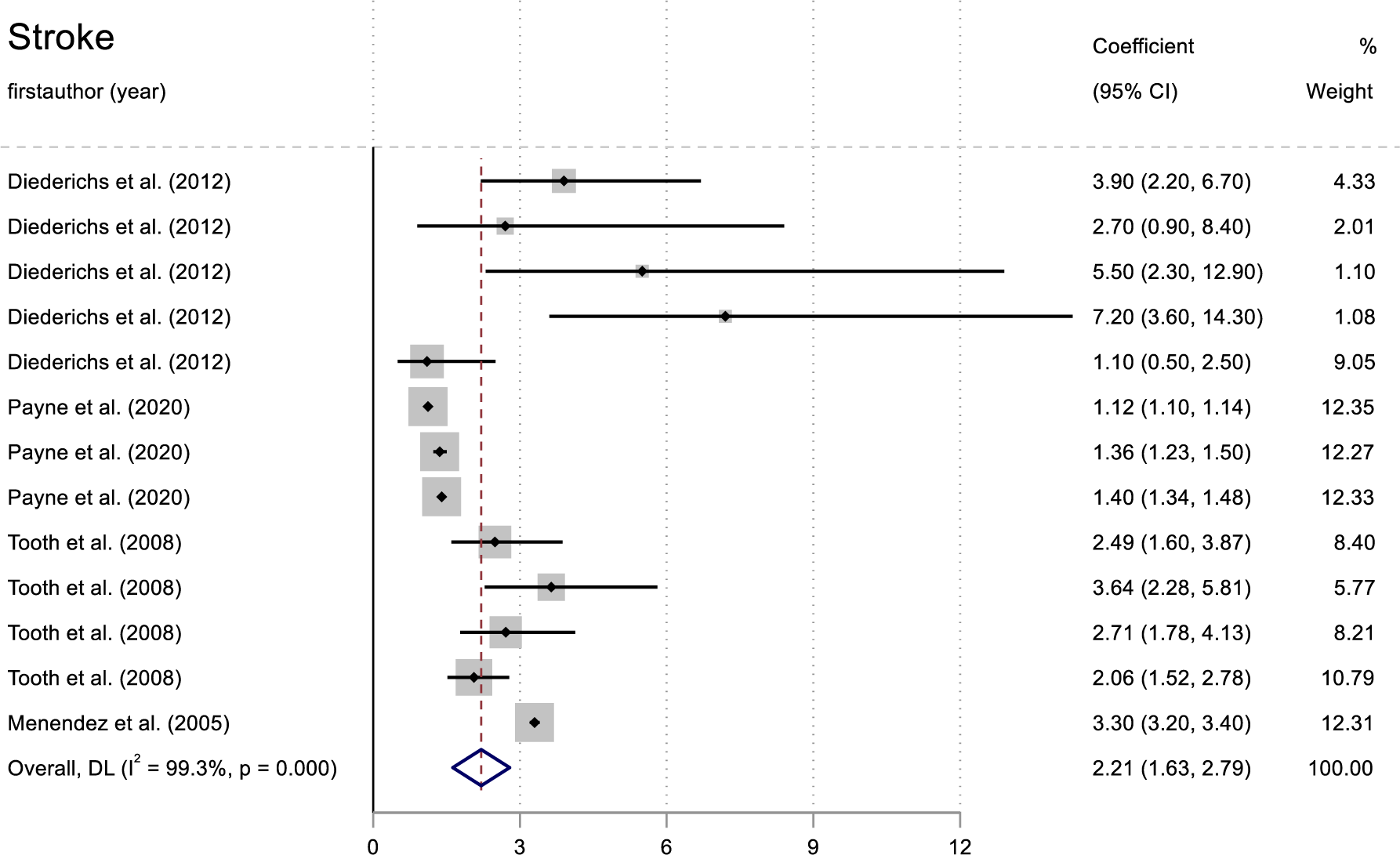
Pooled weight of 16 conditions using random effect meta-analysis at age 63.

## Notes

### Competing Interest Statement

The authors have declared no competing interest.

